# Should I stay or should I go? Observation post-vaccination during the COVID-19 pandemic and the law of unintended consequences

**DOI:** 10.1101/2022.04.05.22273373

**Authors:** Gerard Ingham, Rebecca Kippen

## Abstract

**Background:** Standard practice after all vaccinations in Australia is to observe patients for 15 minutes. During the COVID-19 pandemic, could the risk of contracting and dying from COVID-19 acquired in the waiting room be greater than the risk of dying from post-vaccine anaphylaxis when leaving immediately?

**Methods:** The risks are modelled for a patient aged 70+ years attending for annual influenza vaccination in a typical Australian general practice clinic. The risk of death from anaphylaxis is estimated based on known rates of anaphylaxis shortly after influenza vaccination. The risk of acquiring COVID-19 during a 15-minute wait and then dying from that infection is estimated using the COVID-19 Aerosol Transmission Estimator and COVID-19 Risk Calculator.

**Results:** Other than at times of extremely low COVID-19 prevalence, the risk of death from anaphylaxis for a patient aged 70+ years leaving immediately after influenza vaccine is less than the risk of death from COVID-19 acquired via aerosol transmission during a 15-minute wait. The risk of death from COVID-19 is greatest for the unimmunised and when masks are not worn.

**Conclusions:** A more nuanced approach to advice post-vaccination is recommended that considers current COVID-19 prevalence and virulence, the characteristics of the waiting room, the risk of anaphylaxis, and the patient’s susceptibility to death from COVID-19. There are many circumstances where it would be safer for a patient to leave immediately after vaccination.

*“If I go there will be trouble. And if I stay it will be double”*

*The Clash*

## Introduction

Short-term (15-minute) observation after vaccination is standard clinical practice for all vaccines administered in Australia. This enables prompt management of immediate adverse reactions, particularly anaphylaxis, to reduce the risk of hospitalisation and death (Australian Technical Advisory Group on Immunisation 2018b).

Prior to winter, adults aged 65+ years are strongly recommended to receive an influenza vaccine (Australian Technical Advisory Group on Immunisation 2018a). This year (2022) the incidence of COVID-19 infections is high and expected to increase coming into winter (Clun 2022). There is a risk of acquiring COVID-19 infection while waiting in a shared waiting room after influenza vaccination.

For patients attending for their annual influenza vaccine, will the risk of death from contracting COVID-19 in the waiting area be greater than the risk of death from anaphylaxis? In what circumstances should the current after-care advice be changed to instead instruct patients to leave immediately after their influenza vaccination?

In this paper we explore these questions by modelling the risks for patients aged 70+ years attending a typical general practice for their annual influenza vaccine. We estimate the increased risk of death from anaphylaxis were patients to leave immediately after vaccination versus the increased risk of contracting COVID-19 in the waiting room, and the subsequent risk of dying from it. We model different scenarios considering the impact of the background rate of COVID-19 in the community, patients’ COVID-19 vaccination status, and whether those present are wearing masks.

Approval for the research was granted by the Monash University Human Research Ethics Committee (Project ID 32465)

### The risk of death from anaphylaxis for a patient leaving immediately post-vaccination

Serious adverse reactions after vaccination including anaphylaxis are rare and, with current practice, death almost never occurs. The reported anaphylaxis rate post influenza vaccine is 1.6 cases per million doses (McNeil 2019). For older adults, around 35% of post-vaccination anaphylaxis occurs within 30 minutes of vaccination (McNeil *et al*. 2016). It is likely that patients with reactions within 1–2 minutes of vaccination will still be identified and treated, and many who suffer anaphylaxis after leaving the clinic will be able return in time to obtain medical care. In estimating the increased risk of death from anaphylaxis if patients leave immediately post-vaccination rather than waiting 15 minutes, we hypothesise that half of the patients experiencing anaphylaxis in the first 30 minutes will die resulting in 0.3 deaths per million influenza vaccine doses.

### The risk of contracting COVID-19 in the waiting room post-vaccination

COVID-19 can be transmitted by droplet, fomite, and aerosol spread (World Health Organization 2022). Australian general practices are currently advised to disinfect surfaces and maintain physical distancing in waiting rooms to mitigate against the risk of fomite and droplet spread (Royal Australian College of General Practitioners 2022). Accordingly, to estimate the risk of contracting COVID-19 in the waiting room we used the COVID-19 Aerosol Transmission Estimator (Jimenez and Peng 2022) which models the likelihood of contracting COVID-19 in indoor spaces via aerosol transmission alone. We have used the January 2022 version (3.6.7) that includes an adjustment for the Omicron COVID-19 variant.

The COVID-19 Aerosol Transmission Estimator uses a box model to estimate the concentration of COVID-19 in the indoor air and combines this with the Wells-Riley infection model to estimate the risk of infection (Peng *et al*. 2022). The Wells-Riley infection model is a well-accepted method to quantify risk of infection, initially developed to understand the aerosol transmission of measles and tuberculosis (Riley *et al*. 1978; Riley 2001). In the COVID-19 Aerosol Transmission Estimator, empirical data from known and well-documented COVID-19 transmission events has been used to determine the amount, or ‘quanta’, of aerosolised COVID-19 virus required to cause infection. In other words, the COVID-19 Aerosol Transmission Estimator is a model that has been calibrated using real world data.

Full details of the assumptions used in our calculations are available in Appendix 1. In brief, our typical general practice waiting room ‘box’ was developed by first adopting the recommended waiting room design of 6 waiting room chairs per general practitioner (GP) and 2 metres^2^ per chair = 12m^2^ per GP (Royal Australian College of General Practitioners 2012). Two-thirds of GPs in Australia work in a group practice of 3-10 doctors (Royal Australian College of General Practitioners 2021). We selected 6 doctors as a midway figure, resulting in a waiting room area of 72m^2^. We assumed 2.4 metre ceilings and an absence of filtering of recirculating air or HEPA filters. A ventilation rate of 2h-1 was chosen for the waiting room. General practice clinics are not usually subject to ventilation standards, and many are converted houses. The ventilation figure selected is between that for a house with windows closed and one with windows open.

The risk of acquiring COVID-19 infection is significantly influenced by either past infection or immunisation. To accommodate this, the model permits the inclusion of an estimate of the percentage of the population immune from infection. At the time of our proposed influenza immunisation clinic prior to winter 2022, there is little immunity from COVID-19 due to previous infection in people aged 70+ years in Australia. Background rates of COVID 19 infection in this age group have been low. Protection from infection will mostly be due to immunisation. At the end of March 2022, over 95% of the adult Australian population have received two doses of COVID-19 vaccine and 67% of the eligible population had received a third dose of a COVID-19 vaccine (Australian Government Department of Health 2022b). Most elderly Australians received AstraZeneca vaccine for their first two COVID-19 vaccine doses before receiving an mRNA vaccine as their third (booster) vaccine dose. For this cohort, based on studies by UK public health agencies and international data, expert consensus is that vaccine effectiveness against symptomatic infection with the Omicron variant is 60 % within 3 months of the third vaccine, 40% at 4-6 months after the third vaccine, and only 5% more than 6 months after the second vaccine (UK Health Security Agency 2022). We have therefore selected for our model the fraction of the population immune from infection (not immunity from severe disease or death) as 50%. There may be future scenarios with new COVID variants where population immunity to infection is lower than 50%.

For the influenza immunisation clinic for patients aged 70+ years, two models were created for 6 vaccinators operating with 5-minute appointments. The first model represents current clinical practice of 20 minutes in total in the waiting room, 5 minutes before and 15 minutes after vaccination, with an average of 24 people present. The second model represents altered practice, with 6 patients waiting 5 minutes before the appointment and then leaving immediately after vaccination. The increased risk of contracting COVID-19 with a 15-minute wait post-vaccination (versus no wait post-vaccination) is estimated as the net difference of risk between the two models.

It is not possible to predict either the prevalence of COVID-19 disease in the community or government policy regarding mask requirements at the time of influenza vaccinations this year (2022). To account for this uncertainty, we ran our models for different rates of COVID-19 infectivity prevalence in the community (1%, 0.5%, 0.1%, 0.025%) and for those in the waiting room either all wearing masks or all not wearing masks. The outcomes for the eight modelled scenarios are shown in Figure 1. The risk of contracting COVID-19 varies from a high of 3.31 cases per 1,000 15-minute ‘waits’ (for community infectivity prevalence of 1% and everyone unmasked) to a low of 0.02 per 1,000 (for prevalence of 0.025% and everyone masked).

**Figure 1.**
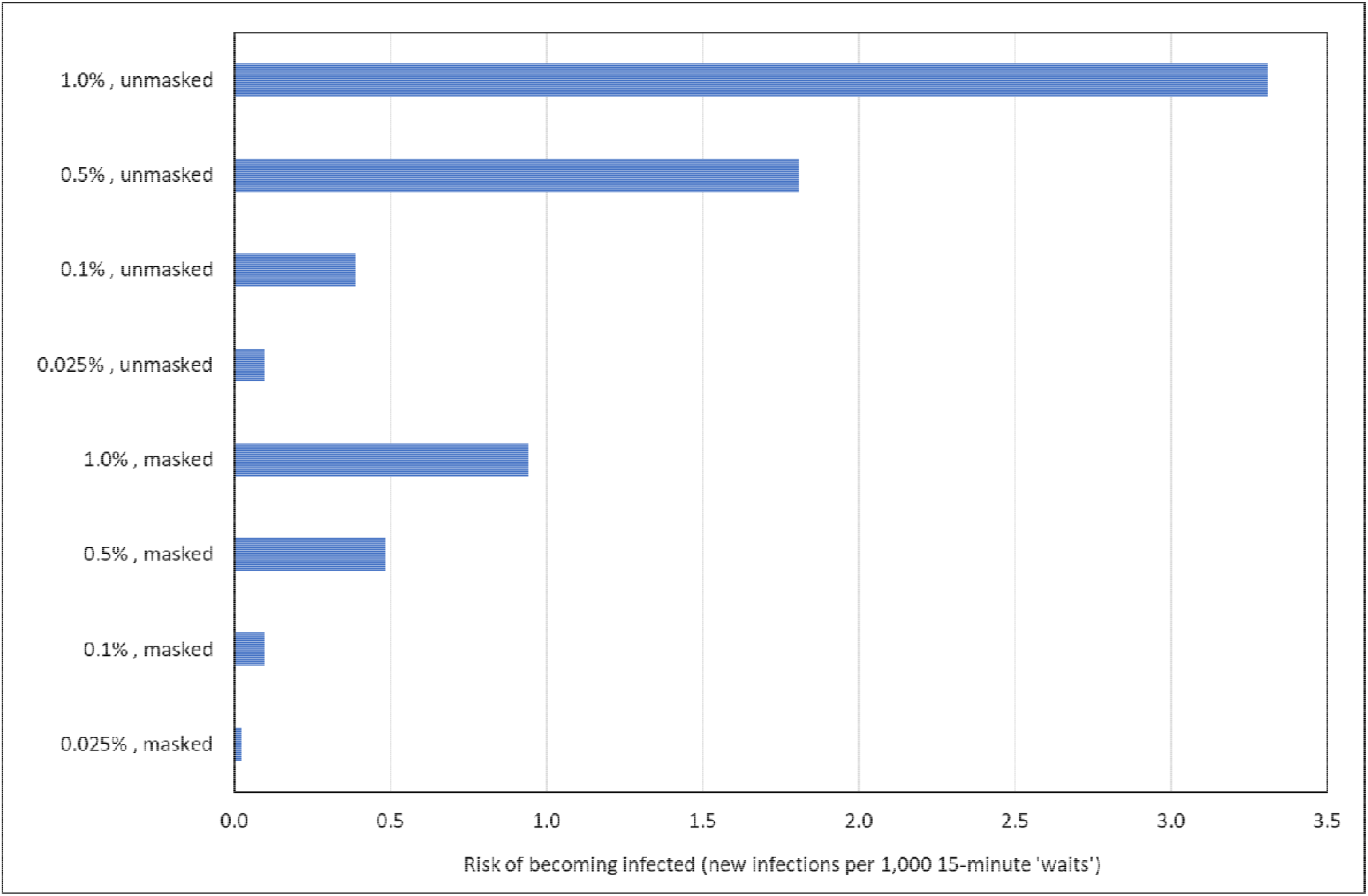
Risk of becoming infected with COVID-19 (new infections per 1,000 15-minute ‘waits’), by selected population infectivity prevalence (1%, 0.5%, 0.1%, 0.025%), and whether patients are masked or unmasked. Calculations are based on the assumptions in Appendix 1 inputted to the COVID-19 Aerosol Transmission Estimator (Jimenez and Peng 2022)

### The risk of death from COVID-19 infection contracted in a waiting room

If an infection is contracted in the waiting room, the risk of dying (the case fatality rate) is influenced by many factors of which the COVID-19 variant, and patient age, sex and vaccination status are among the most significant. To estimate the COVID-19 case fatality rate we used the summary chart produced by the Immunisation Coalition based on their COVID-19 Risk Calculator for Australian conditions in early 2022 that includes adjustment for the Omicron variant (Immunisation Coalition 2022). For the age group 70+ years, the COVID-19 case fatality rate varies from a high of 36.2 deaths per thousand COVID-19 cases for men who are not immunised against COVID-19, to 1.3 per thousand for women with a recent booster (third dose). Rates are presented in Figure 2.

**Figure 2.**
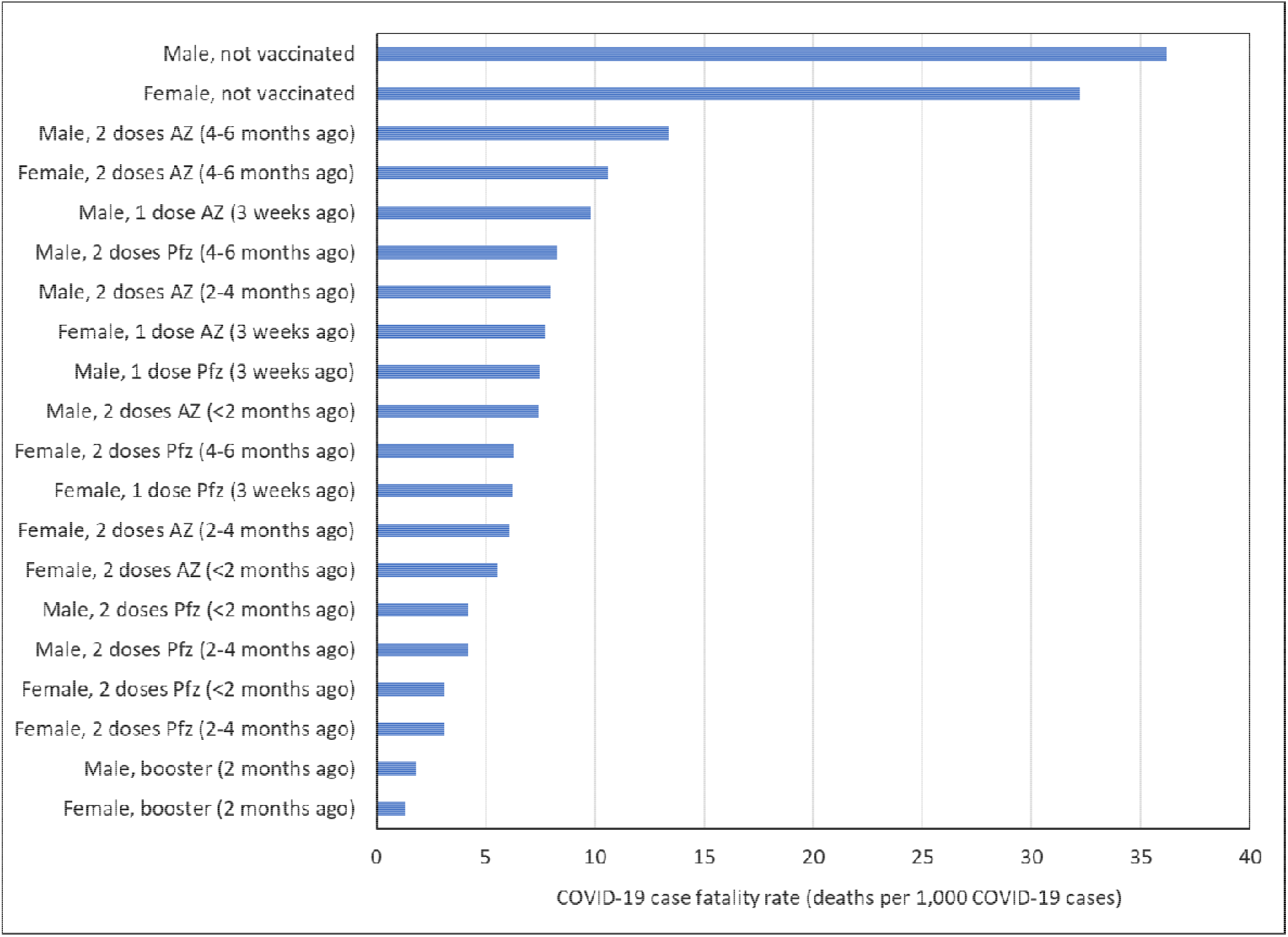
COVID-19 case fatality rate (deaths per 1,000 cases), Australian population aged 70+ years by sex and COVID-19 vaccination status. Rates are drawn from the COVID-19 Risk Calculator. (Immunisation Coalition 2022). AZ=AstraZeneca COVID-19 vaccine; Pfz=Pfizer COVID-19 vaccine; booster=third dose of either Pfizer or Moderna COVID-19 vaccine

### When should a patient aged 70+ years be advised to leave immediately after influenza vaccination?

The combined risk of a patient aged 70+ years contracting COVID-19 during a 15-minute wait post-vaccination and subsequently dying from the infection is displayed in Table 1 under different scenarios, calculated by multiplying the rates in Figure 1 by the rates in Figure 2. Scenarios is which it would be safer to leave (risk of anaphylaxis of 0.3 per million is less than the risk of contracting and dying from COVID-19) are unshaded.

**Table 1.**
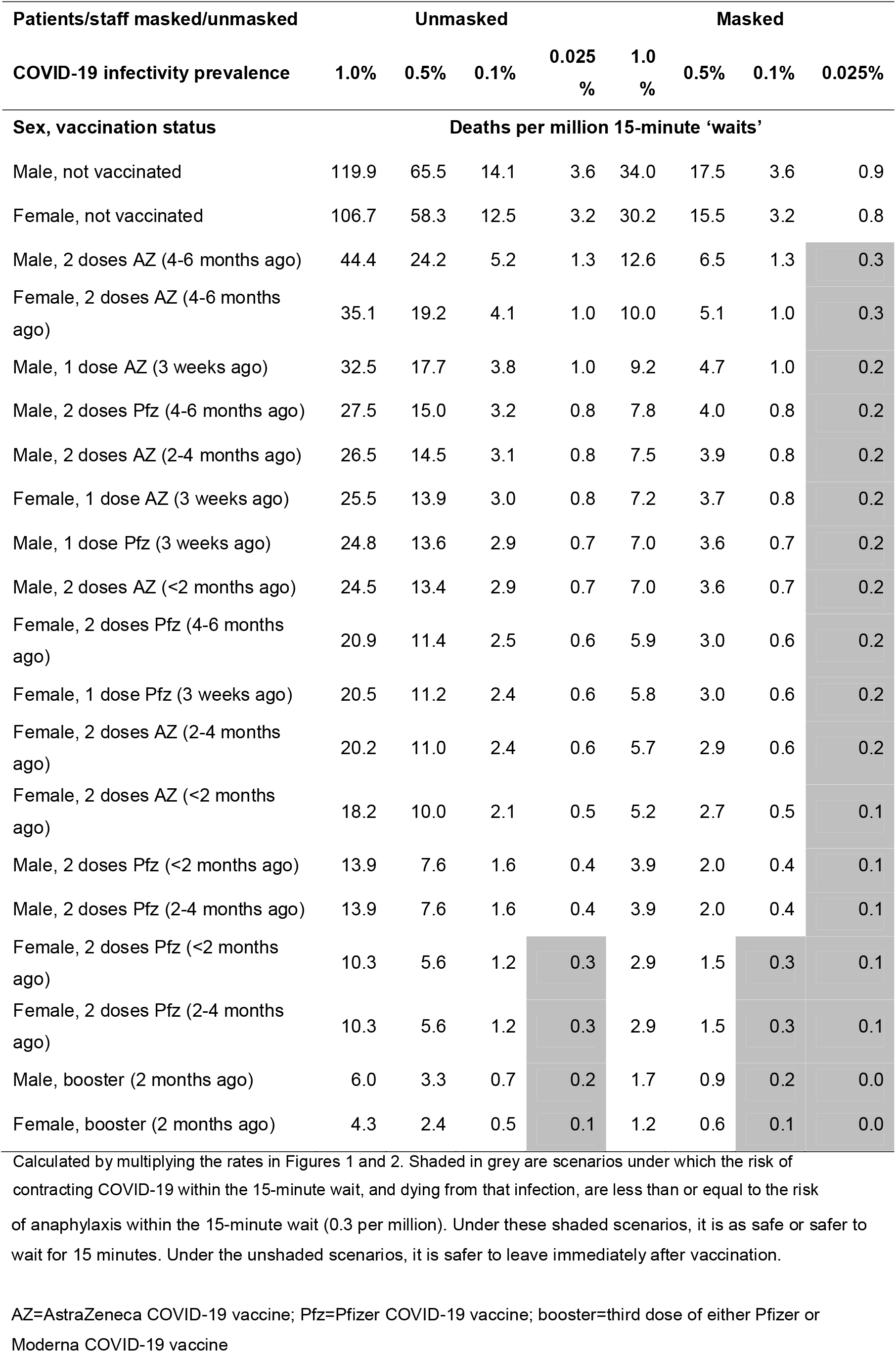
Risk of becoming infected with COVID-19 and dying from that infection (deaths per million 15-minutes ‘waits’), Australian population aged 70+ years, by sex, COVID-19 vaccination status, selected population infectivity prevalence, and whether patients are masked or unmasked Patients/staff masked/unmasked Unmasked Masked

When community COVID-19 infectivity prevalence is 0.5% (1 in 200) or greater, it is safer for all those aged 70+ years to leave immediately after influenza vaccination, no matter their COVID-19 vaccination status, or whether masked or unmasked. This is because the risk of contracting COVID-19 in the 15-minute wait and dying from that infection is greater than 0.3 per million and thus outweighs the risk of anaphylaxis from influenza vaccine within those 15 minutes.

At the other end of the scale, if community COVID-19 infectivity prevalence is extremely low - 0.025%, or 1 in 4,000 - then it is safer for most people aged 70+ years to stay for 15 minutes after influenza vaccination (COVID-19 risk is equal or lower than anaphylaxis risk), but only if they are immunised. Even then, failure to wear a mask would render those with sub-optimal immunisation status unsafe.

## Discussion

The risks of either suffering anaphylaxis from a vaccination or dying from COVID-19 contracted while being observed post-vaccination are both tiny; they are both far outweighed by the benefits of immunisation in preventing serious illness and death. The question considered in this paper is whether the benefit of staying for 15 minutes observation post-influenza vaccination – in order to treat any cases of post-vaccination anaphylaxis - is outweighed by the risk of contracting COVID-19 within those 15 minutes and dying from that infection. For the population aged 70+ years, we have shown that unless community prevalence of COVID-19 is extremely low the risk of anaphylaxis after influenza vaccination is indeed outweighed by the risk of COVID-19, and it would be safer for patients to leave immediately after vaccination.

Our analysis is relatively simple and has been designed conservatively. Our model presumes physical distancing can be maintained and all droplet and fomite transmission is prevented. Our highest modelled COVID-19 prevalence infectivity rate of 1% is not extreme. At the height of the most recent ‘Omicron wave’ in mid-January 2022 there were an estimated 766,000 active cases in Australia (Australian Government Department of Health 2022a), a prevalence of 3% of Australia’s population. The lowest modelled infectivity rate of 0.025% (1 in 4,000) is unlikely to be reached until after winter 2022.

Vaccination in Australia occurs in a wide variety of settings, from small pharmacies through to drive-through clinics. We modelled an intermediate risk setting - a medium-sized general practice. Our model has not included risk factors other than age, immunisation status, mask wearing, and background community infectivity prevalence. The risk of death from anaphylaxis if leaving earlier than 15 minutes will be greater for those with a history of anaphylaxis. The risk of death from COVID-19 acquired waiting 15 minutes longer in the waiting room will be higher for those with comorbidities like diabetes or asthma.

In determining whether to advise a patient to stay or go after a vaccination, a clinician needs to consider the risk of death from anaphylaxis from that vaccine were the patient to leave, the risk of death were they to contract COVID-19, and the risk of acquiring COVID-19 a*t that time* and *in their waiting room*.

In April 2020, at the commencement of the COVID-19 pandemic, when droplet spread was considered the major mode of COVID-19 transmission and prior to the availability of COVID-19 immunisations, the Australian Technical Advisory Group on Immunisation (ATAGI) advised only 5-minutes observation was required after vaccination if physical distancing could not be achieved in the waiting room (Australian Technical Advisory Group on Immunisation 2020). Our model included physical distancing of at least 1.5 metres in the waiting room (1 patient per 3m^2^) and excluded the risk of droplet and fomite spread. Our findings indicate that there are circumstances where aerosol transmission alone is enough to advise against the patient remaining in the waiting room.

We propose that more nuanced advice that considers changing COVID-19 prevalence and virulence be given to vaccination providers about vaccination after-care. Patients with a history of anaphylaxis may be safer to be observed after vaccination, but elderly and particularly unimmunised patients should be instructed to leave unless the background infectivity rate of COVID-19 is very low. Patients should continue to wear masks when attending indoor vaccination clinics and physical distancing should be maintained.

A more detailed analysis could include risks of acquiring other infections, such as influenza, in the waiting room. There are health and economic benefits from the more efficient operation of vaccination clinics when the 15-minute-wait requirement is removed (The Chief Medical Officers of the UK and lead Deputy Chief Medical Officers for vaccines 2021). Disregarding COVID-19, there may still be circumstances where the net benefit of a 15-minute wait after vaccination does not exist.

From a public health perspective our example illustrates the operation of what has been termed the ‘law of unintended consequences’. Policies may be designed with good intention—in this case to ensure prompt management of anaphylaxis—but may unintentionally have adverse outcomes that outweigh any benefit (Newman *et al*. 2003; Turcotte-Tremblay *et al*. 2021)

## Supporting information

data file for table

## Data Availability

All data produced in the present work are contained in the manuscript and appendix

## Data Availability Statement

The data that support this study are available in the article and accompanying online supplementary material.

