## Appendix 1 for "Should I stay or should I go? Observation post-vaccination during the COVID-19 pandemic and the law of unintended consequences"

### Appendix 1: Should I stay or should I go? Observation post-vaccination during the COVID-19 pandemic and the law of unintended consequences

#### Background

We have used the COVID-19 Aerosol Transmission Estimator version 3.6.7 date 27 Jan 2022 available at https://docs.google.com/spreadsheets/d/16K1OQkLD4BjgBdO8ePj6ytf-RpPMlJ6aXFg3PrIQBbQ/edit#gid=519189277

This version is based on 100% Omicron variant

The output we are obtaining from this calculator is the COVID -19 infection rate in the waiting room. We have not used this COVID -19 Aerosol Transmission calculator to estimate the death rate from COVID acquired in the waiting room. We are instead using the Immunisation Coalition's COVID-19 risk calculator to calculate the likelihood of death once infection occurs. This is because the Immunisation Coalition's calculator allows stratification according to immunisation status and has been adapted for the current Australian situation of various combinations of primary immunisations and booster vaccines

As there are no general practice waiting room models in the Aerosol Transmission Estimator, we selected and edited the recommended "Class" model which is the master spreadsheet that can be used for any scenario. This file lists the assumptions we used in the 'Class' model and the justification for the inputs chosen.

#### Calculating the Number of Additional COVID cases resulting from waiting 15 minutes

To calculate the number of additional cases of COVID caused by requiring a patient to wait we created a model for:

Current Practice (C) - patient remains for 15-minutes post vaccination
Altered Practice (A) - patient leaves immediately post vaccination

The absolute difference in COVID cases generated in the waiting room is the difference between the outcomes for the 2 models

C - A = no of additional cases of COVID in waiting room for waiting 15 minutes

#### Scenarios Modelled

<u>Prevalence</u>
High prevalence of COVID in the community - infectivity rate of 1.0%
Medium prevalence of COVD in the community - infectivity rate of 0.5
Low prevalence of COVID in the community – infectivity rate of 0.1
Very low prevalence COVID in the community – infectivity rate of 0.025

<u>Mask-wearing</u>
And wearing mask or not wearing mask

<u>Consequently, we have outcomes (C-A) for 8 scenarios:</u>
HighMask, MedMask, LowMask, VeryLowMask
HighNomask, MediumNoMask, LowNoMask, VeryLowNoMask

### The following parameters were inserted the calculator

#### Environmental Parameters

Our waiting room size of $72m^2$ is based on a pre-pandemic waiting room design and an estimate of the average size of a general practice as follows:

- 6 waiting room chairs are recommended per doctor and 2 $metres^2$ per chair = $12m^2$ per doctor Reference: Starting a Medical Practice Guide available from https://www.racgp.org.au/FSDEDEV/media/documents/Running%20a%20practice/Practice%20resources/Management%20toolkit/Starting-a-mediacl-practice.pdf

- ⅔ of GPs in Australia work in group practice between 3-10 doctors. We selected 6 as a midway figure. Reference: State of Nation Report RACGP 2021 available from https://www.racgp.org.au/health-of-the-nation/chapter-2-general-practice-access/2-2-gp-workforce

The height of the waiting room selected is 2.4 metres = average ceiling height in Australia (volume is $173m^3$)

We accepted the standard parameters of 0.95atm, 20C, 50% humidity, 415 ppm background CO2 outdoors

Duration of the event was either 20 minutes (C) or 5 minutes (A). This is because in both situations we estimated a patient will be in the waiting room for 5 minutes waiting for the doctor and transiting the waiting room after the consultation. Current practice (C) includes an additional 15 minutes in the waiting room whereas in altered practice (A) the patient leaves immediately after the consultation

For the number of repetitions of the event we have selected 1000, consequently the absolute outputs from our calculation are recorded as events per 1000 immunisations.

For ventilation we estimate a rate of 2h-1 being greater than that of a house but less than that of a house with windows open. Many GP surgeries in Australia are in converted houses or if commercially built do not have specific ventilation standards like other health facilities.

We accepted default parameters of Decay rate of virus 0.62 h-1, Deposition to surfaces 0.3 h-1

We presume the application of no additional control measures (e.g. filtering of recirculated air or HEPA air cleaner) as these are not currently broadly in operation in Australia.

#### Parameters related to people and activity in the room

The duration of the event (patient time in the waiting room) and the number of people in the room is calculated for the two different scenarios as below and is based on a 5-minute booking schedule for influenza vaccine which we consider to be a conservative estimate for a vaccination clinic.

Current Practice (C)
Duration of event = 20 minutes. In addition to 15 minutes waiting post vaccination, we have allowed 5 minutes for pre-appointment waiting and time exiting to the reception. We have not included time in the vaccination room.

For a 5-minute appointment schedule we estimate that the waiting room will contain one person waiting for vaccination and three people post vaccination = 4 persons per doctor = **24** people in the waiting room. This is likely a conservative estimate

Altered Practice (A): patient leaves immediately post vaccination
Duration of event is 5 minutes = time for pre-appointment waiting and exiting to reception

Number of people in waiting room = 1 person waiting for vaccination per doctor = **6** people in the waiting room

For infective people we have inserted 1. The Aerosol Transmission calculator provides outputs based on a conditional result for one event (conditional on one infective person being present) or an absolute rate based on disease prevalence (probability of being infective). We are reporting the latter, so our selection of 1 is recommended to obtain this output. This is further explained in this video https://youtu.be/2acW7ruCRKM?t=2179

We have selected the fraction of the population immune from infection as 50%. This figure is for immunity from infection (not severe disease or death). The risk of acquiring COVID-19 infection is significantly influenced by either past infection or immunisation. At the time of our proposed influenza immunisation clinic prior to winter 2022, there is little immunity from COVID-19 due to previous infection in people aged 70+ in Australia. Background rates of COVID 19 infection in this age group have been low. Protection from infection will mostly be due to immunisation. At the end of March 2022, over 95% of the adult Australian population have received two doses of COVID-19 vaccine and 67% of the eligible population had received a booster dose of a COVID-19 vaccine. Most elderly Australians received AstraZeneca vaccine for their first two doses before receiving a booster with an mRNA vaccine. For this cohort, based on studies by UK public health agencies and international data, expert consensus estimates vaccine effectiveness against symptomatic infection with the Omicron variant to be 60 % within 3 months of a booster dose, 40% 4-6 months after a booster dose, and only 5% more than 6 months after a second dose UK Health Security

Agency We have therefore selected for our model the fraction of the population immune from infection (not immunity from severe disease or death) as 50%. There may be future scenarios with new COVID variants where population immunity to infection is lower than 50%. Therefore, we consider a 50% assumption to be reasonable for our model and on the conservative side

Our scenario is for people over 70 attending for influenza vaccine so we have used the breathing rate for 71-80 year olds: the breathing rate is 12.9 $m^3$/day = 0.5735 $m^3$ /hr

We assumed a basic quanta emission rate of 4.7 which is the factor for people resting and speaking as per the “Read Me” section of the calculator. We considered the alternative option of people sitting still and not speaking but did not considered it to be truly representative of a medical waiting room behaviour. People do converse with each other or with their partners or family members attending with them. Also, people called into the doctor's office often respond verbally at that time.

We assumed a 2.5 quanta enhancement for the Omicron variant – the recommended input – as this is the predominant strain in Australia currently (March 2022)

We assumed no enhancement (a value of 1) due to vocalisation (singing or shouting) or activities (exercise)

We modelled either 100% mask wearing or zero mask wearing

We applied a 50% mask efficiency for both inhalation and exhalation as this is the recommended default value. This is explained further in the ‘ReadMe’ section of the calculator. Based on our observation of mask-wearing compliance in waiting rooms this is similarly a generous calculation.

#### Parameters related to the COVID 19 disease

The disease prevalence has been run for 4 scenarios: VeryLow (0.025%) , Low (0.1%), Medium (0.5%) and High (1%). Now that COVID has become widespread in Australia, a rate of 1 in 4000 people is a very low level and unlikely to be achieved prior to this (2022) winter. The Omicron wave in January Australia had a prevalence of at least 3%, or likely more due to testing services being overwhelmed at the time. An infectivity rate of 1/3 of active infections is a conservative assumption. According to the 'ReadMe' section in the calculator, NYC in 2020 had an infectivity rate of 2.3%. All scenarios assume the rate of infective people in the community will be the same as our patients aged 70+ years in the waiting room. It may be that infective people are more likely to be in the doctor's waiting room. Anecdotally there were cases of people attending COVID immunisations after attending exposure sites due to being worried about their health and this would increase the percentage of infective people in the waiting room. Alternatively, if screening procedures are present on entry to the practice to exclude infective people, the rate may be lower.

We have entered a hospitalization rate of 0% and a death rate of 0% as we are instead using the COVID 19 risk calculator for these outputs

### Results

As above we are not reporting either conditional results (conditional on one person being infected) or hospitalisation or death outcomes (we are using COVID 19 risk calculator for these)

From this calculator, we are only reporting the Absolute result for events that are repeated multiple times - the number of COVID cases arising. The outputs are cases per 1000 vaccinations as we have modelled 1000 repeats of each vaccination event

The outputs were calculated for the 8 scenarios by considering number of cases in current practice (C) minus the number of cases in altered practice for (A)

We obtained the following results:

| **6.0 DOCTOR GP MODEL** | | | | | | |
|---|---|---|---|---|---|---|
| | Current Practice Mask | Altered Practice Mask | Difference | Current Practice No Mask | Altered Practice No Mask | Difference |
| High (1.0) | 0.94 | 0 | 0.94 | 3.33 | 0.01 | 3.32 |
| Medium (0.5) | 0.48 | 0 | 0.48 | 1.82 | 0.01 | 1.81 |
| Low (0.1) | 0.1 | 0 | 0.1 | 0.39 | 0 | 0.39 |
| Very Low (0.025) | 0.02 | 0 | 0.02 | 0.1 | 0 | 0.1 |
